# Proton pump inhibitors are detrimental to overall survival of patients with glioblastoma multiforme: results from a nationwide real-world evidence database

**DOI:** 10.1101/2024.01.28.24301899

**Authors:** Michael P. Castro, Jameson Quinn, Asher Wasserman, Alaa Awawda, Zachariah D. Cole, Mark A. Shapiro, Timothy J. Stuhlmiller, Santosh Kesari

## Abstract

**Background:** Aldehyde dehydrogenase-1A1 (ALDH1A1) is a primary metabolic enzyme impacting outcome of chemotherapy, including temozolomide, the standard-of-care (SOC) for glioblastoma multiforme (GBM). High expression of ALDH1A1 is associated with poor prognosis in multiple cancers. Proton pump inhibitors (PPIs) are often prescribed to manage corticosteroid-induced gastrointestinal toxicity but were recently identified as strong inducers of ALDH1A1, suggesting a negative impact on survival.

**Methods:** Real-world data on GBM patients was annotated from electronic medical records (EMR) according to the prospective observational study, XCELSIOR (NCT03793088). Patients with *IDH1/2* mutations were excluded. Causal effects on survival were analyzed using a multivariate, time-varying Cox Proportional Hazard (CPH) model with stratifications including *MGMT* methylation status, age, sex, duration of corticosteroid use, extent of resection, starting SOC, and PPI use.

**Results:** EMR data from 554 GBM patients across 225 cancer centers was collected, with 72% of patients receiving care from academic medical centers. Patients treated with PPIs had numerically lower median overall survival (mOS) and 2-year OS rates in the total population and across most strata, with the greatest difference for MGMT-methylated patients (mOS 29.2 mo vs. 40.1 mo). In a time-varying multivariate CPH analysis of the above strata, PPIs caused an adverse effect on survival (HR 1.67 [95% CI 1.15-2.44], p=0.007).

**Conclusions:** Evidence from a nationwide cancer registry has suggested PPIs have a strong detrimental effect on OS for GBM patients, particularly those with *MGMT* promoter methylation. This suggests PPIs should be avoided for prophylactic management of gastrointestinal toxicity in patients with GBM receiving SOC chemoradiotherapy.

**Key Points:**

- Nationwide glioblastoma study suggests hazardous effect of proton pump inhibitors
- Discretion advised when prescribing proton pump inhibitors to glioblastoma patients
- Prophylactic proton pump inhibitor use to limit chemo toxicity may increase risk

**Importance of the Study:** Recent molecular evidence suggests off-target activity of proton pump inhibitors (PPIs) may counteract the activity of alkylating chemotherapy in glioblastoma clinical care. Utilizing a time-varying cox proportional hazard model and abstracted clinical data from medical records according to a nationwide observational research protocol, we found PPIs were significantly associated with greater risk of death, independent of corticosteroid use. Importantly, the detrimental effect was most impactful for patients with methylated MGMT promoters who gain the most benefit from standard-of-care temozolomide treatment and experience the best outcomes.

## Introduction

Glioblastoma multiforme (GBM) is one of the most aggressive types of cancer with a median overall survival (mOS) of 14.6 months and 5-year survival of ∼5%.^1^ Current estimates of OS from CBTRUS in 2022 reported the mOS to be 8 months after implementing the updated 2021 WHO diagnostic criteria requiring *IDH1/2*-wild type status.^2,3^ The standard-of-care (SOC) treatment involves maximal surgical resection, concurrent temozolomide (TMZ) and external beam radiation therapy, followed by maintenance TMZ with tumor-treating fields. MGMT is a key protein involved in the reversal of DNA methylation resulting from TMZ alkylation. In addition to being a crucial predictive biomarker for TMZ benefit, *MGMT* promoter methylation is the strongest known prognostic biomarker for OS in GBM.

Proton pump inhibitors (PPIs) are liberally prescribed in neurosurgery and neuro-oncology as prophylaxis against GI bleeding and dexamethasone-induced gastropathy. PPIs are conventionally thought to be benign in the context of malignant disease, or even potentially beneficial through reversal of the acidic tumor microenvironment (TME). However, several epidemiologic studies have showed small but significant increases in mortality among patients taking PPIs for many conditions, including cancer, but these studies had potential confounding that could not be resolved with the available data.^4-6^

A key to understanding these previous observations is the recent finding that proton pump inhibitors, such as omeprazole and pantoprazole, are potent inducers of ALDH1A1.^9-11^ Extensive literature demonstrates ALDH1A1 is a major mediator of therapy resistance and is associated with poor prognosis across a wide variety of malignancies.^10-17^ In GBM patients, ALDH1A1 expression above the mean causes TMZ and radiation resistance and is strongly associated with reduced survival, while knockdown of ALDH1A1 expression restores sensitivity to chemotherapy and radiation therapy.^18-20^ ALDH1A1 is also a mediator of resistance to EGFR blockade in GBM and non-small cell lung cancer, and appears to activate HIF1A, a major driver of radiation resistance.^21-23^ ALDH1A1 detoxifies alkylating agents and serves as a key antioxidant, reversing lipid peroxidation and repairing etheno-DNA adducts.^24^ Lipid peroxidation leads to cell death through a caspase-independent mechanism known as ferroptosis, thought to play a key role in the outcome of GBM.^25-31^ In addition to enhancing oxidative stress resistance and maintaining REDOX homeostasis that confer chemotherapy and radiation failure, ALDH1A1 catalyzes the conversion of retinaldehyde to retinoic acid resulting in the stemness phenotype that causes perpetual tumor re-population.^32^

Those and other recently reported observations about PPIs suggest that they are not benign or neutral agents. For example, PPIs can alter the TME to promote immunosuppression by enhancing MDSC infiltration and interfere with the T-cell trafficking necessary for the efficacy of PD-L1 inhibitors.^33,34^ PPIs also enhance *YAP1* oncogene activation and alter the gut microbiome in a way that increases the conversion of colonic adenoma to carcinoma.^35^

By contrast, some investigators have tried to show that PPI may have antitumor effects.^36^ The working hypothesis is that by blocking vacuolar-ATPases (proton pumps *ATP6V0A1 and ATP6V0A2*), PPIs deactivate the pH inversion that acidifies the tumor microenvironment and raises intracellular pH. In theory, this would diminish the invasive phenotype, promote apoptosis, and enhance chemotherapy sensitivity.^37,38^ In laboratory experiments, inhibition of tumor invasiveness by PPIs has been reported for GBM.^39^ However, the clinical relevance of these observations is uncertain since the concentrations employed against cell lines are significantly above the C_max_ values achievable in patients.^3,40-43^ Thus far, no randomized trials have been conducted to address these issues in glioma patients. Here we report on the survival outcomes of patients from a national real-world database of GBM patients whose complete longitudinal cancer histories and medication use are known.

## Materials and Methods

### Observational protocol

Patients consented to XCELSIOR (<u>NCT03793088</u>), a central IRB-approved, nationwide, ambispective observational pan-cancer registry, permitting retrospective data collection, and prospective follow-up. With patient authorization, medical records — including both structured elements and unstructured document images — were gathered from all available sites of clinical care for each patient. A median of 2,404 clinical records per patient were gathered from over 8,800 individual locations, with a median of 46 encounter locations per patient. Structured and unstructured data was collected from diverse locations including neuro-oncology clinics, cancer centers, and radiology centers in addition to outpatient labs, infusion centers, primary care, and family medicine clinics (Figure S1). Unstructured text from clinic narratives and digitized PDF images with relevant keywords were utilized as source documents for annotation in an electronic database. Annotated data were source-verified, merged with structured data elements, and mapped to coding systems such as SNOMED, LOINC, and RxNorm to generate standardized longitudinal histories for aggregate analysis.^44^ Accurate dates of diagnosis were abstracted from pathology reports. Patient identity verification permitted determination of accurate death dates and overall survival calculations.

### Cohort identification and definitions

Patients were identified by a reported diagnosis of Glioblastoma multiforme, WHO grade IV on pathology reports. The dataset includes some patients harboring pathogenic *IDH1* or *IDH2* mutations. Since the WHO 2021 diagnostic criteria redefined glioblastoma as IDH-wild type and because IDH mutations are prognostic for longer OS, patients with known *IDH1* or *IDH2* mutation were excluded from analysis.

Strata were split as follows: age (<60 or ≥60 years), sex (male/female), *MGMT* promoter methylation status (methylated, unmethylated, unknown). Extent of surgical resection (total, partial, none) was determined by review of clinic notes and radiology reports and was coded to “total” if found to be total resection, gross total resection, or near-total resection; was coded to “partial” if noted to be partial resection or subtotal resection; was coded to “none” if biopsy-only was performed; in all cases, this was restricted to a 150-day observation window around GBM diagnosis (30 days before and 120 days after diagnosis). Patients were stratified based on whether they started SOC treatment (received temozolomide and/or radiation therapy) in the same 150 day observation window around diagnosis.

### Statistical Analysis

The cutoff date for analysis was August 1, 2023. Kaplan-Meier curves (with log-rank statistics) were produced for various splits: *MGMT* methylation status (methylated, unmethylated, or unknown); PPI use in the 150 day observation window around diagnosis; PPI use within *MGMT* subcategories; days of corticosteroid use in the 150 day observation window diagnosis (<15, 15-60, >60); age at diagnosis (<=60, >60); and assigned sex at birth. OS is reported from date of diagnosis to date of death, or the date last known alive, determined by the most recent clinic note or medication date in the EMR.

To determine the cutpoints for continuous variables of corticosteroid duration and age at diagnosis, we performed a Cox proportional-hazards (CPH) analysis with a spline expansion of the continuous terms. We used a B-spline expansion of order 4 with breakpoints determined by the data quantiles. The cutpoints in the age and steroid variables were chosen to match the changes in the effect of the covariates on survival as seen in the resulting spline fits.

We conducted a statistical analysis using the CPH model incorporating time-varying binary variables^45^ for two key covariates: PPI usage (1 only if a patient has received post-diagnosis PPI) and “steroid dependency” (1 if a patient has had 60+ consecutive days on corticosteroids post-diagnosis). Fixed covariates included in the model were extent of resection, age at diagnosis, and sex. This analysis excluded patients with unknown *MGMT* methylation status and those who did not start SOC treatment because this population exhibited greater heterogeneity, thereby amplifying the potential impact of selection bias. This resulted in 273 patients for analysis.

Sensitivity analyses were also performed by repeating the principal time-varying CPH analysis, varying the time to “steroid dependency” (15 or 30 days, rather than 60). To eliminate the potential influence of corticosteroid utilization spanning the 15-60 day range, we conducted an auxiliary analysis involving one time-varying variable with three potential states: ‘0’ (baseline) if patients abstained from PPI use post-diagnosis, ‘1’ if they initiated PPI use without concurrent corticosteroid use in the 15 days prior, and ‘2’ if PPI initiation coincided with corticosteroid use within the preceding 15 days. This analysis used the same population and fixed covariates as above. It should be noted that a drawback of this approach is that the effect estimate of interest, pertaining to group ‘1’, establishes a lower boundary for the impact within a subgroup with lower total corticosteroid utilization, a factor unobservable at the point of diagnosis.

## Results

Between April 2019 and April 2023, a total of 605 patients with a pathology-proven diagnosis of GBM were enrolled. Fifty-one (51) patients had *IDH1/2*-mutant tumors that were transformed from WHO Grade III anaplastic astrocytoma or low grade gliomas, or diagnosed as *IDH*-mutant GBM by 2016 WHO criteria. As expected, *IDH* mutation was significantly associated with longer mOS (58.9 months vs. 21.5 months, Figure S2). For subsequent analyses, patients with *IDH*-mutant GBM were excluded, leaving a total of 554 patients in the analysis cohort.

These 554 patients resided in 48 US states and received oncology care from a total of 225 cancer centers or health systems (Figure 1B,C). In this dataset, 49% of patients received oncology care exclusively at academic medical centers, 28% exclusively at community hospitals/health systems, and 23% received care at both types of centers (Figure 1D). Altogether, 72% of patients received care or consults from at least one academic medical center.

**Figure 1:**
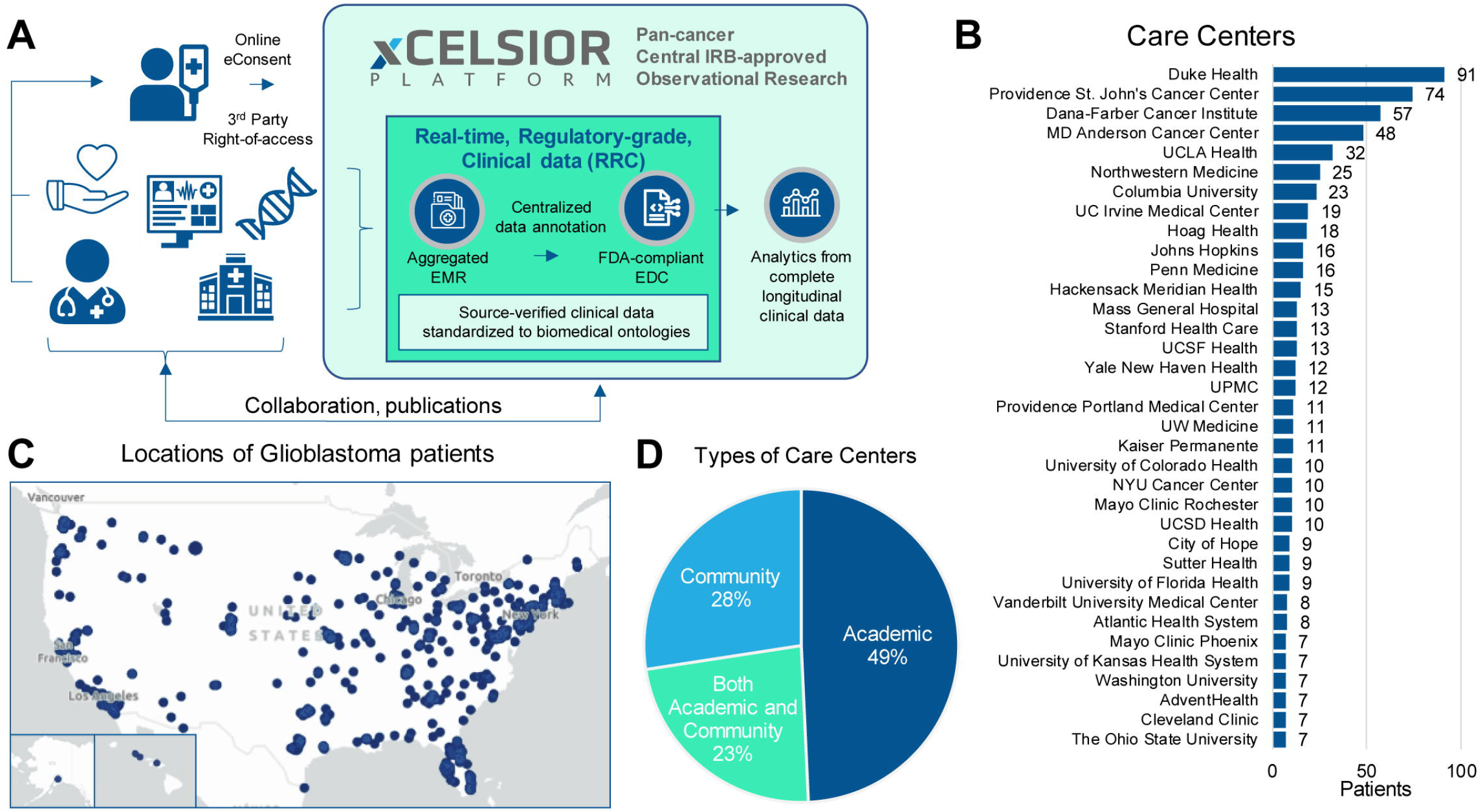
Platform summary and glioblastoma patient treatment sites (A) Summary diagram of the XCELSIOR real world evidence platform. Patients with cancer or suspected cancer consent electronically (eConsent) to the XCELSIOR master observational research protocol. Through HIPAA 3rd party right-of-access, medical records are aggregated from all sites of care, inclusive of EMR, radiology, and genomics results. Data is annotated in a central 21 CFR Part 11-compliant electronic data capture (EDC) system and coded to OMOP-based ontologies. Standardized data is used for analysis. (B) Top 35 care centers by number of patients who were treated by those sites. Same patient may be counted in more than one site. (C) Home residences of patients with glioblastoma used in the analysis. Size of bubble is proportional to number of patients by zip code. (D) Distribution of the types of care centers visited by patients.

The median date of diagnosis among GBM patients in the analysis cohort was November 13, 2020 and median age at diagnosis was 55 years. Median follow-up from diagnosis was 13.8 months (interquartile range 8.0-22.3 months) and 265 deaths had occurred by the analysis cutoff date.

Among 554 patients, 286 (51%) had been exposed to PPIs at any point and 215 (39%) started PPIs during the 150-day observation window for analysis (30 days prior to diagnosis to 120 days after diagnosis). Primary covariates of age, sex, *MGMT* promoter methylation status, extent of surgical resection on diagnosis, duration of corticosteroid use, starting SOC, and type of care site were balanced between patients exposed or not exposed to PPIs (Table 1).

**Table 1:** Primary clinical features of cohort.

| Covariate | Covariate level | Total population | PPI | no PPI |
| --- | --- | --- | --- | --- |
| PPI exposure | Exposed | 215 (39%) | 215 (100%) | - |
| PPI exposure | Not exposed | 339 (61%) | - | 339 (100%) |
| Sex at birth | Male | 359 (65%) | 141 (66%) | 218 (64%) |
| Sex at birth | Female | 195 (35%) | 74 (34%) | 121 (36%) |
| Age category | < 60 years | 359 (65%) | 133 (62%) | 226 (67%) |
| Age category | ≥ 60 years | 195 (35%) | 82 (38%) | 113 (33%) |
| MGMT status | Methylated | 163 (29%) | 74 (34%) | 89 (26%) |
| MGMT status | Unmethylated | 328 (59%) | 130 (60%) | 198 (58%) |
| MGMT status | Unknown | 63 (11%) | 11 (5%) | 52 (15%) |
| Resection status | Total | 232 (42%) | 87 (40%) | 145 (43%) |
| Resection status | Partial | 224 (40%) | 90 (42%) | 134 (40%) |
| Resection status | None | 98 (18%) | 38 (18%) | 60 (18%) |
| Steroid category | < 15 days | 240 (43%) | 43 (20%) | 197 (58%) |
| Steroid category | 15 - 59 days | 146 (26%) | 82 (38%) | 64 (19%) |
| Steroid category | ≥ 60 days | 168 (30%) | 90 (42%) | 78 (23%) |
| SOC treatment | Started | 507 (92%) | 207 (96%) | 300 (88%) |
| SOC treatment | Did not start | 47 (8%) | 8 (4%) | 39 (12%) |
| Care site | Academic only | 273 (49%) | 109 (51%) | 164 (48%) |
| Care site | Community only | 152 (27%) | 46 (21%) | 106 (31%) |
| Care site | Both academic and community | 129 (23%) | 60 (28%) | 69 (20%) |

Kaplan-Meier (K-M) curves were generated and median overall survival (mOS) and 2-year OS were calculated for subgroups across the whole population (Figure 2 and Table 2). In the overall population, exposure to PPI resulted in a numerical but non-significant reduction in mOS (20.3 months exposed vs. 21.4 months not exposed) and 2-year survival (42% exposed vs. 46% not exposed). Subgroup analysis, stratified on PPI use within the 150 day observation window, suggested that PPI use was associated with reduced mOS and 2-year OS particularly in the population with *MGMT* methylated status and in the one with age < 60 years old, but these findings did not reach statistical significance (Figure 3). These univariate analyses are useful for understanding the key strata and validating the expected effects of known prognostic features, but because of potential differences in these prognostic features between PPI exposed and unexposed patients our principal analysis was designed to control for these differences and other potential sources of confounding or bias in real world data.

**Table 2:** Median OS and 2-year OS by primary strata.

|  |  | mOS, months (95% CI) |  |  | 2-year OS (95% CI) |  |  |
| --- | --- | --- | --- | --- | --- | --- | --- |
| Feature |  | Total Population | PPI | No PPI | Total Population | PPI | No PPI |
| Sex |  |  |  |  |  |  |  |
|  | Male | 20.3 (18.0-21.7) | 19.8 (16.6-22.1) | 21.0 (17.7-23.8) | 40% (34%-47%) | 37% (26%-47%) | 42% (34%-50%) |
|  | Female | 26.4 (20.3-32.7) | 25.4 (16.5-53.5) | 26.4 (20.3-33.0) | 53% (44%-61%) | 55% (39%-68%) | 53% (42%-63%) |
| Age |  |  |  |  |  |  |  |
|  | < 60 years | 22.4 (20.7-27.6) | 20.7 (18.6-26.3) | 24.5 (21.1-31.8) | 48% (42%-55%) | 44% (33%-55%) | 50% (42%-58%) |
|  | ≥ 60 years | 17.3 (14.1-20.1) | 16.6 (12.6-25.8) | 17.3 (14.0-21.3) | 37% (28%-46%) | 39% (25%-54%) | 35% (24%-46%) |
| MGMT status |  |  |  |  |  |  |  |
|  | Methylated | 39.7 (28.9-50.7) | 29.2 (19.4-53.5) | 40.1 (28.3-53.5) | 68% (58%-76%) | 61% (45%-74%) | 73% (60%-82%) |
|  | Unmethylated | 17.7 (16.2-19.8) | 18.5 (15.6-20.5) | 17.7 (15.1-20.3) | 31% (24%-37%) | 27% (17%-39%) | 31% (23%-39%) |
|  | Unknown | 26.8 (16.0-42.4) | 26.8 (2.9-29.5) | 18.9 (16.0-59.8) | 53% (37%-66%) | 42% (7%-76%) | 51% (34%-66%) |
| Resection status |  |  |  |  |  |  |  |
|  | Total | 25.4 (20.9-29.2) | 22.5 (17.2-26.3) | 26.4 (21.2-33.0) | 53% (44%-6%) | 48% (33%-61%) | 55% (45%-64%) |
|  | Partial | 19.0 (16.2-21.4) | 19.0 (15.6-21.7) | 19.0 (15.2-21.7) | 37% (29%-50%) | 33% (2%-50%) | 39% (29%-50%) |
|  | None | 20.0 (14.0-27.6) | 20.3 (11.4-29.5) | 18.6 (14.0-27.6) | 42% (30%-50%) | 50% (28%-68%) | 39% (24%-54%) |
| Steroid use |  |  |  |  |  |  |  |
|  | < 15 days | 21.7 (19.1-25.7) | 20.6 (12.6-24.2) | 21.7 (19.1-27.6) | 45% (37%-52%) | 33% (16%-51%) | 47% (38%-55%) |
|  | 15-59 days | 19.4 (17.2-26.1) | 18.6 (16.6-25.8) | 20.1 (15.2-32.3) | 43% (32%-54%) | 40% (25%-54%) | 47% (31%-62%) |
|  | ≥ 60 days | 21.4 (17.6-28.9) | 20.9 (15.5-29.5) | 21.4 (16.8-28.9) | 45% (35%-54%) | 47% (32%-60%) | 44% (30%-56%) |
| SOC |  |  |  |  |  |  |  |
|  | Started | 21.4 (19.7-24.3) | 20.5 (17.7-25.4) | 21.7 (19.7-26.2) | 45% (40%-51%) | 44% (34%-53%) | 46% (39%-53%) |
|  | Did not start | 14.2 (11.1-27.6) | 10.5 (3.5-12.5) | 14.5 (11.6-33.7) | 36% (19%-53%) | 0% | 44% (24%-62%) |

**Figure 2:**
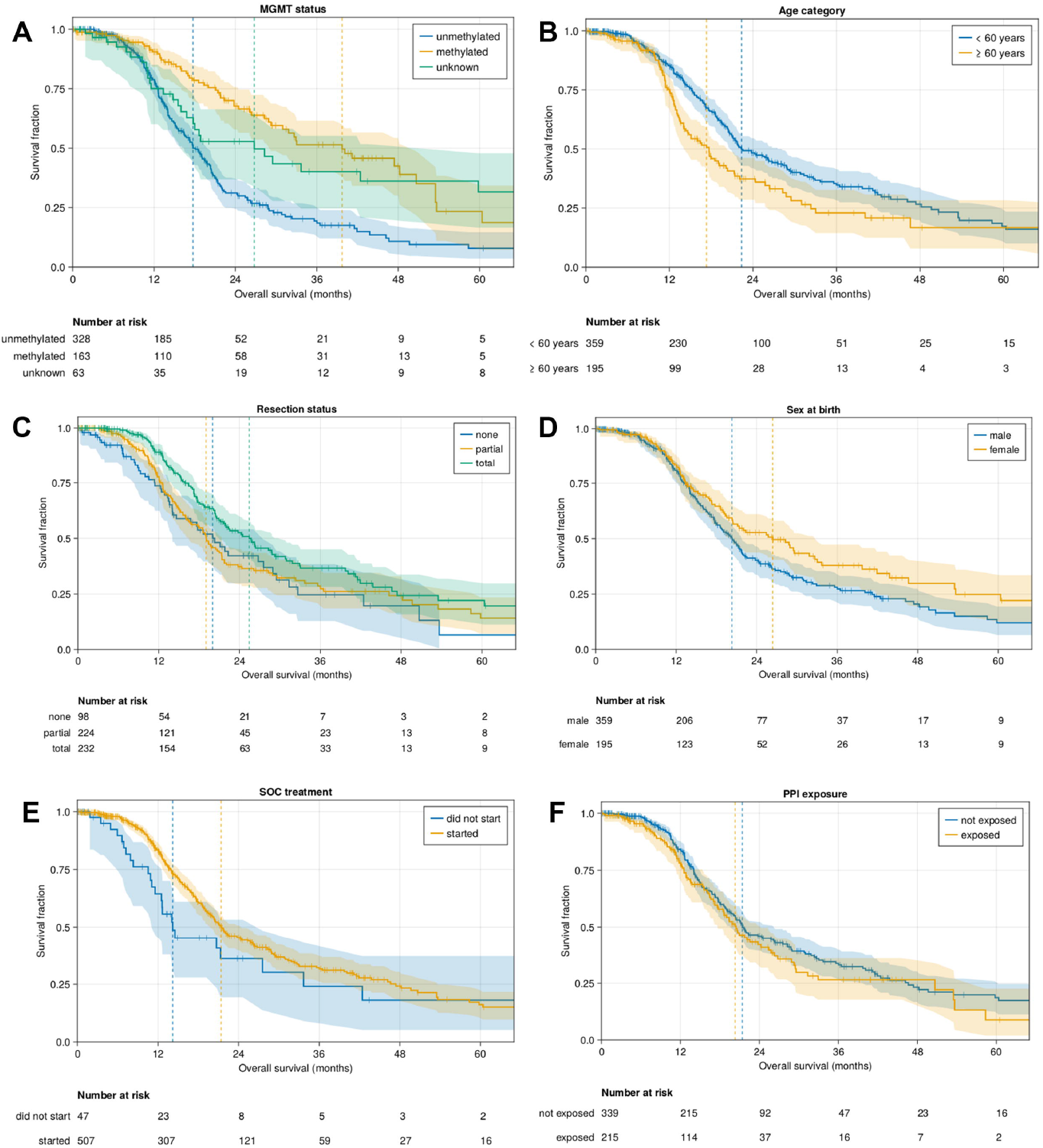
Baseline clinical strata with impacts on overall survival Kaplan-Meier curves for primary clinical strata: (A) MGMT promoter methylation status, (B) Age, (C) Sex, (D) extent of resection, (E) starting standard-of-care (SOC) with 120 days of diagnosis, and (F) starting proton pump inhibitors (PPIs) within 150 day observation window around diagnosis.

**Figure 3:**
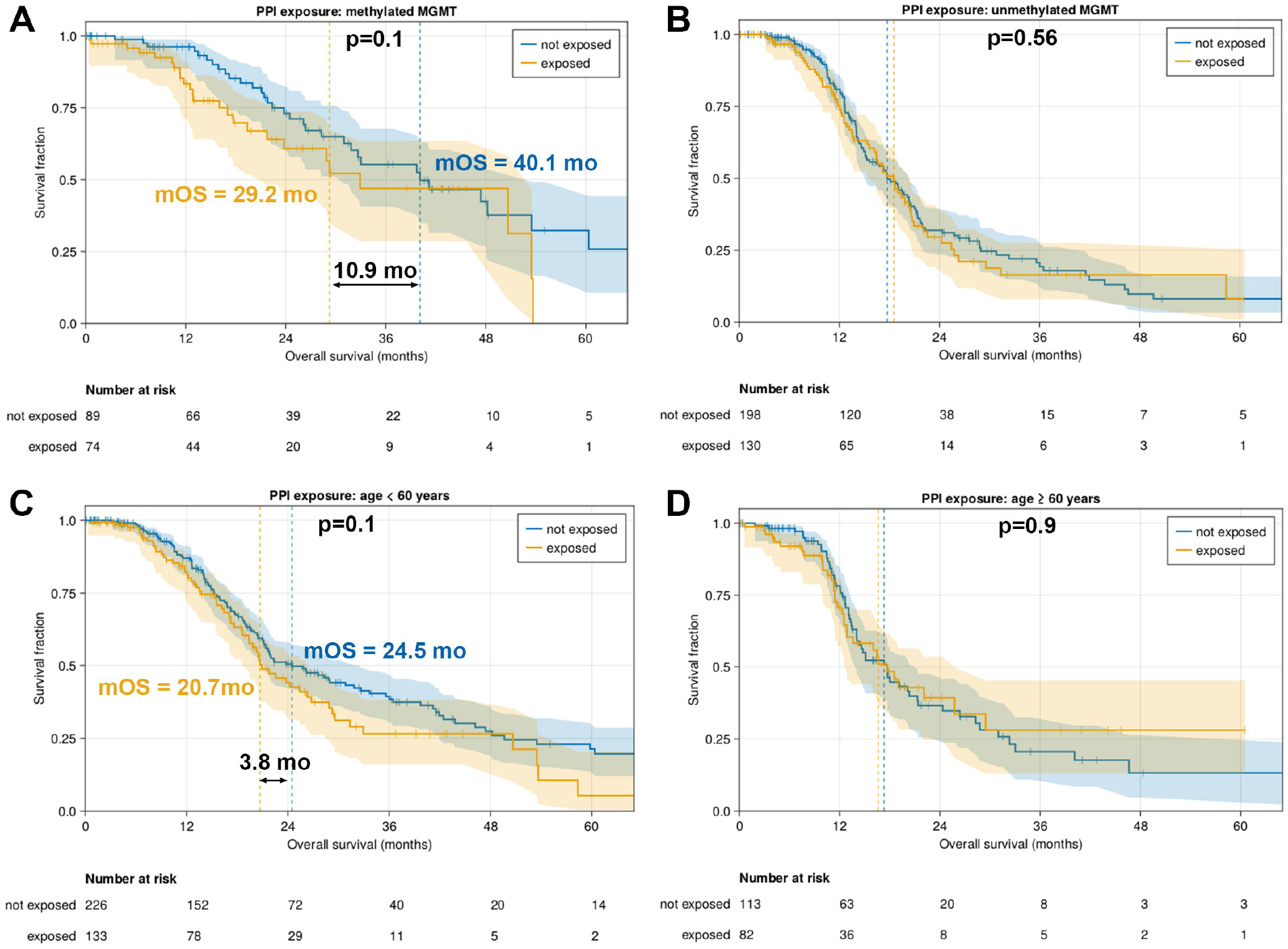
Proton pump inhibitor use reduces OS of patients with methylated MGMT and age < 60 Kaplan-Meier plots for patients exposed or not exposed to PPIs during the 150-day observation window for subpopulations with (A) methylated MGMT promoter, (B) unmethylated MGMT promoter, (C) age <60 years, (D) Age > or = 60. PPI, proton pump inhibitor. mOS, median overall survival. Logrank p-values are provided in each panel.

PPIs are often prescribed prophylactically with corticosteroids or in response to adverse effects from corticosteroid use. Accordingly, PPI exposure and corticosteroid exposure showed the greatest correlation of all covariates (R^2^ = 0.37). Spline modeling of corticosteroid duration on hazard ratio indicated corticosteroid use between 15 and 60 days during the 150-day observation window generated the greatest risk of death (Figure S3), suggesting prolonged use of corticosteroids during this phase of disease was detrimental. To resolve potential confounding between corticosteroids and PPI use and to account for PPI use that occurs more than 120 days after diagnosis, we employed a multivariate Cox proportional-hazards model with time-varying binary variables for PPI use and corticosteroid dependency. 273 patients were used in this analysis (see Methods), creating 692 patient-periods. In this multivariate, time-varying CPH analysis, PPI use showed a significantly increased risk of death (HR 1.67 [1.15 to 2.44], *p*=0.007). As expected, methylated *MGMT* status showed a significantly reduced risk of death (HR 0.34 [0.19 to 0.61], *p*<0.001). However, much of the benefit of MGMT methylation was abrogated by PPI use (Figure 4), consistent with the hypothesis that the action of PPIs is through a reduction in the beneficial effects of chemoradiotherapy.

**Figure 4:**
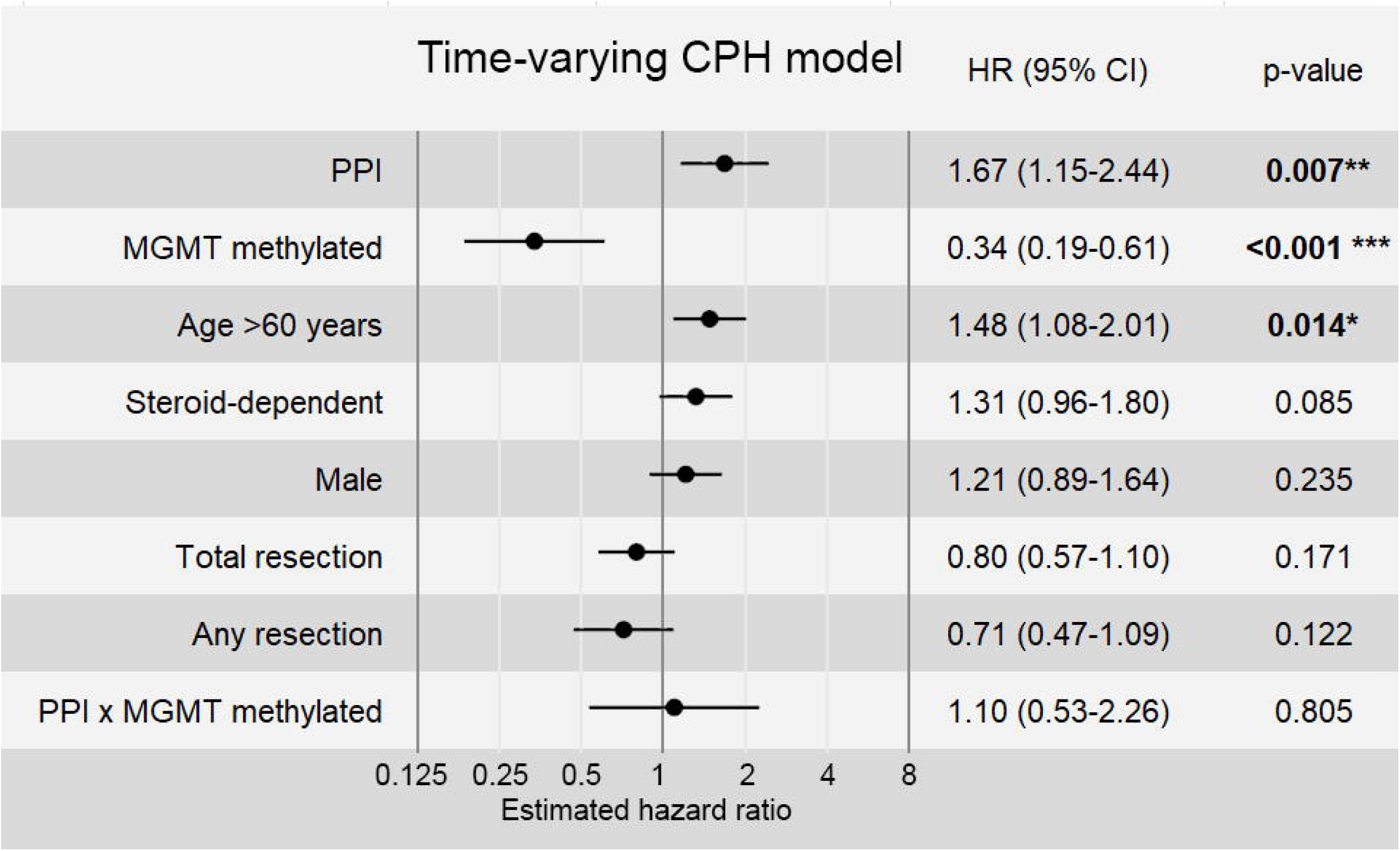
Proton pump inhibitor use is hazardous to survival by time-varying Cox Proportional Hazard model Cox proportional-hazards (CPH) model incorporating time-varying binary variables for two covariates: Proton pump inhibitor (PPI) usage and “steroid dependency” defined as patients requiring 60+ days of consecutive use of corticosteroids at any point in their treatment.

Corticosteroid dependence was of borderline significance in this model, with an increased risk (HR 1.31, *p*=0.085). Sensitivity checks on this model, varying the time to steroid dependency (15 or 30 days) showed the significance of coefficients was robust; both had significant *p* values (P_null_[|z|*>|z|]<0.05) for the PPI and *MGMT*-methylated coefficients. In fact, these sensitivity analyses also brought the *p* value for corticosteroid use below 0.05 (d=15:*p*=0.034; d=30:*p*=0.020), though care should be taken with these values as they do not include any multiple-comparisons corrections.

Since post-baseline variables can be problematic when treated as a baseline covariates, we performed a landmark analysis starting 120 days from diagnosis in the time-varying CPH model. This generated the same trends as observed in the primary analysis, (Figure S4). To eliminate the potential influence of temporally proximal corticosteroid use with PPIs, we conducted an auxiliary time-varying analysis. Results for this auxiliary analysis show a significantly higher hazard for patients that started PPIs without concomitant corticosteroid use compared to patients that were not exposed to PPIs, supporting the primary analysis (Figure S5).

## Discussion

Utilizing a unique nationwide real-world dataset, we found evidence that PPI use places GBM patients at increased risk of death. Patients whose tumors show *MGMT* promoter methylation displayed the greatest hazard from PPI use. Since this is the population which gains the most benefit from TMZ, this is strongly consistent with the hypothesis that PPIs disrupt the efficacy of alkylating chemotherapy. Accordingly, the CPH model suggested that PPI use appeared to diminish the survival benefit of *MGMT* methylation.

Corticosteroid use also has been tied to worse outcomes in GBM.^46,47^ But despite the emphasis on minimizing the use of corticosteroids in neuro-oncology, patients who are unable to have complete tumor resection often require prolonged corticosteroids to manage vasogenic edema. Leaky vasculature caused by the cancer is commonly exacerbated by treatment effect, thus linking extent of surgery and dexamethasone use with diminished survival. Since PPI use often accompanies corticosteroids for the purpose of GI prophylaxis, it is challenging to differentiate the effect of PPI use from the deleterious impacts of corticosteroids, especially as the need for dexamethasone is co-mingled with other risk factors for diminished survival outcome. Notably, the *MGMT* promoter contains two glucocorticoid receptor response elements, thus linking TMZ resistance to dexamethasone use.^48^ Other deleterious effects of dexamethasone on infection risk, lymphopenia, metabolic disturbances, thromboembolism risk, muscle wasting, and diminished performance status may also contribute to diminished survival.^49,50^ Nevertheless, in this cohort of patients, the deleterious impact of corticosteroid use by multivariate analysis was less than half of the impact of PPI use.

This study is not the first to show that PPI use is associated with reduced survival. A recent epidemiological study replicated previous research showing that PPI prescription was strongly associated with all-cause and cause-specific mortality.^4^ However, the authors downplayed their and others’ findings of increased mortality in lung cancer, mesothelioma, breast cancer, liver cancer, prostate cancer, and gastric cancer, among others, because “a plausible causal mechanism” was lacking. However a compelling causal mechanism has emerged linking ALDH1A1 to therapeutic resistance from oxidative stress resistance and promotion of the cancer stem cell phenotype.

What distinguishes this study from previous epidemiological studies linking PPI use with increased mortality is the access to complete longitudinal patient records and a careful analysis designed to control for the principal potential sources of bias and confounding, such as immortal time bias. To reduce the risk of immortal time bias, we limited our group assignments in the Kaplan-Meier plots to observations made in a 150-day window around diagnosis, and employed this same time frame as the threshold for considering patients as recipients of SOC treatments for their inclusion in the CPH analyses. Sensitivity checks suggest any remaining immortal time bias is negligible.

A second concern with previous epidemiological studies is around the most efficient use of the available data. When creating Kaplan-Meier plots, each patient must be permanently assigned to just one group, requiring one to disregard PPI and corticosteroid steroid usage outside the observation window. The time-varying Cox proportional-hazards analyses presented herein address this issue by accounting for PPI and/or steroid use at any point in time.

The third consideration pertains to the issue of confounding factors. We were particularly concerned about the potential for confounding through corticosteroid dependency. That is, a more severe underlying disease condition could not only directly contribute to higher mortality rates but also induce steroid dependency, potentially resulting in gastric symptoms and subsequently increased PPI usage. Accounting for steroid dependency should mitigate this confounding effect.

For the reasons mentioned above, we believe that the primary time-varying Cox proportional-hazards analysis strikes the best balance, offering the greatest statistical power for detecting a genuine causal effect of PPIs while also being reasonably robust against confounding due to steroid dependency.

In the absence of a randomized controlled trial large enough to permit subgroup analysis, we employed a logical statistical approach to address confounding variables. Significant heterogeneity exists in the presentation and treatment of GBM and we acknowledge that the median followup is short, but we felt the clinical impact of this finding necessitates rapid communication. In the future, we intend to incorporate other relevant features into the model such as performance status and tumor size to further understand the population most at-risk from prophylactic PPI use.

If the estimated hazard ratio for PPI use were an exact estimate of the causal impact, a crude calculation would suggest that of the over 12,000 yearly newly-diagnosed GBM patients in the US, reduced use of PPIs might collectively save nearly 1,500 life-years per year. These results urge caution in the use of PPIs for managing acute upper gastrointestinal bleeding and to avoid prophylactic use of PPIs. The data from this real-world study suggests alternatives to PPIs should be considered whenever possible for GBM patients, particularly among those for whom relatively favorable outcomes are anticipated.

## Funding

No external funding was received for this work.

## Authorship

Conceptualization: MC, MAS; Supervision: TJS, SK, MAS; Methodology, Data Analysis, and Figure Preparation: JQ, AW, TJS; Data curation: AA, ZDC, TJS; Manuscript Writing: All authors; Final approval of manuscript: All authors

## Conflict of Interest Statements

**MPC**: Employment – Cellworks; Stock and Other Ownership Interests - Bugworks; DelMar Pharmaceuticals; Neurovigil; Honoraria - Guardant Health; Consulting or Advisory Role – Omicure; Speakers’ Bureau - Guardant Health; Research Funding - Exact Sciences

**JQ, AW, AA, ZDC**, and **TJS**: Employment – xCures; Stock and Other Ownership Interests - xCures

**MAS**: Employment – xCures; Leadership – xCures; Stock and Other Ownership Interests - xCures; Consulting or Advisory Role - BioNTech; Marinus Pharmaceuticals; Patents, Royalties, Other Intellectual Property - Various patents on software for AI-based clinical decision support technology

**SK**: Stock and Other Ownership Interests - xCures; Honoraria - Jubilant Biosys; Pyramid Biosciences; Consulting or Advisory Role - Biocept; iCAD; xCures; Research Funding - AIVITA Biomedical, Inc. (Inst); Bayer (Inst); Biocept (Inst); Blue Earth Diagnostics (Inst); Boehringer Ingelheim (Inst); Boston Biomedical (Inst); Caris MPI (Inst); cns pharmaceuticals (Inst); EpicentRx (Inst); Lilly (Inst); Oblato (Inst); Orbus therapeutics (Inst)

## Acknowledgements

We would like to acknowledge Al Musella, Marty Tenenbaum, Burt Nabors, Nicholas Blondin, Ekokobe Fonkem, Fabio Iwamoto, Eric Wong, Charles Redfern, and Kamalesh Sankhala.

## Data Availability

Data from XCELSIOR (NCT03793088) is available to academic and government researchers free of charge under license from xCures, Inc. Contact for more information.

## Notes

### Author Declarations

Genetic Alliance Central IRB gave ethical approval for this work

