## Supplemental material for "Proton pump inhibitors are detrimental to overall survival of patients with glioblastoma multiforme: results from a nationwide real-world evidence database"

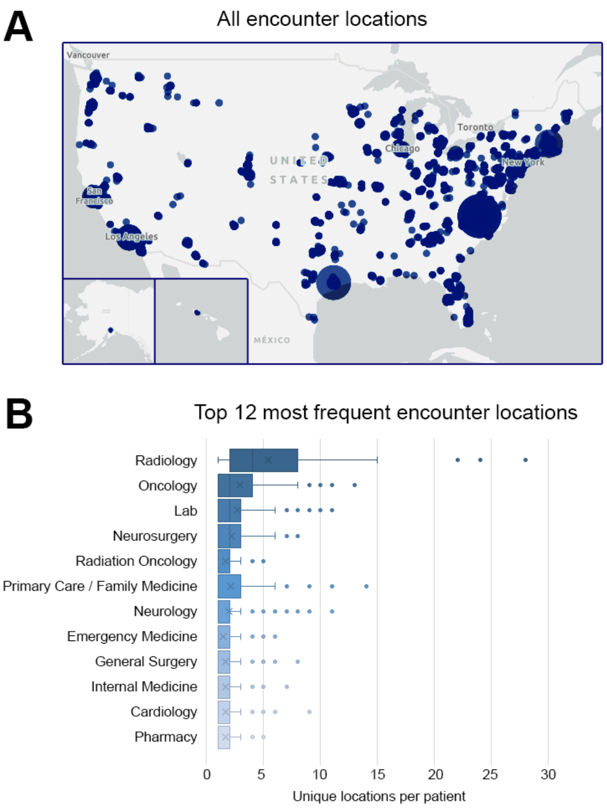


C

| Encounter Location Type | Average unique locations per patient |
|---|---|
| RADIOLOGY | 5.4 |
| ONCOLOGY | 2.9 |
| LAB | 2.7 |
| NEUROSURGERY | 2.2 |
| RADIATION ONCOLOGY | 1.7 |
| PRIMARY CARE/FAMILY MEDICINE | 2.1 |
| NEUROLOGY | 2.0 |
| EMERGENCY MEDICINE | 1.4 |
| GENERAL SURGERY | 1.7 |
| INTERNAL MEDICINE | 1.6 |
| CARDIOLOGY | 1.7 |
| PHARMACY | 1.7 |

**Figure S1: Depth of clinical data collected for patients**

(A) Locations of care centers where records have been gathered for glioblastoma patients. Size of bubble is proportional to number of care locations by zip code. (B) Number of unique locations per patient, stratified by type of encounter location where medical records have been collected. Top 12 most frequent location types across the entire cohort are displayed. (C) Average unique locations per patient for the 12 most frequent encounter location types.

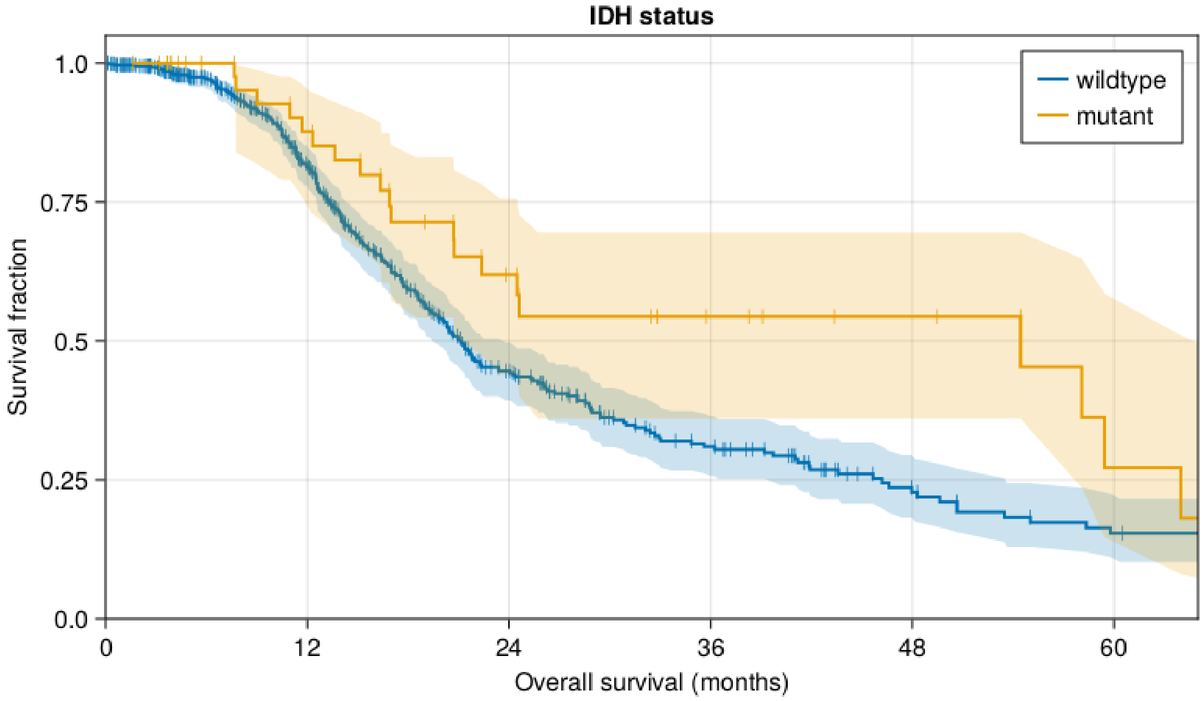


**Number at risk**

| | | | | | | |
|---|---|---|---|---|---|---|
| wildtype | 554 | 329 | 128 | 63 | 29 | 18 |
| mutant | 51 | 36 | 19 | 12 | 8 | 5 |

**Figure S2: *IDH1/2* mutation is associated with longer overall survival**

Kaplan-Meier curve of all patients with pathologically-defined glioblastoma multiforme, stratified on pathogenic *IDH1/2* mutation.

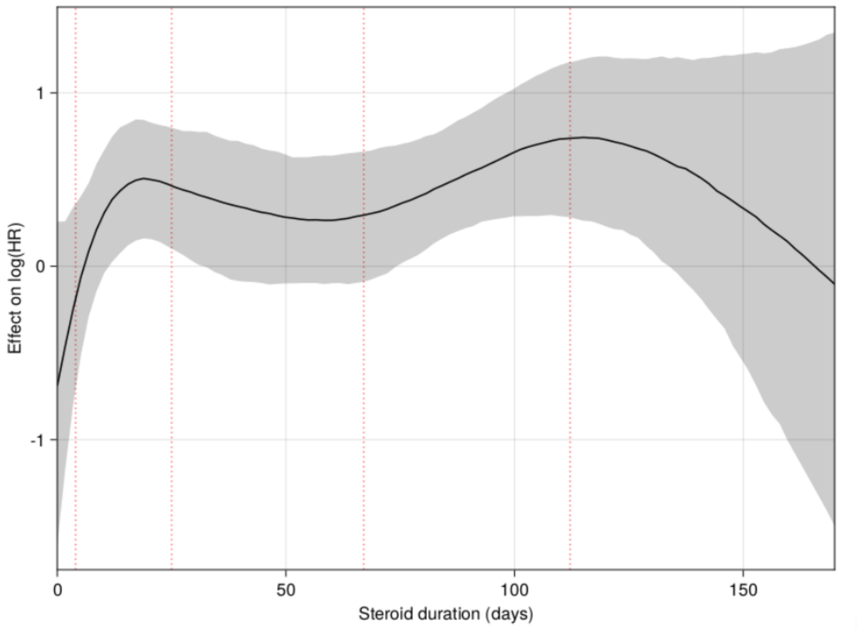


**Figure S3: Steroid use greater than 15 days is associated with greater hazard of death**

Baseline cox proportion hazard modeling of glioblastoma multiforme patients varied by duration of corticosteroid use in a 150-day window around diagnosis.

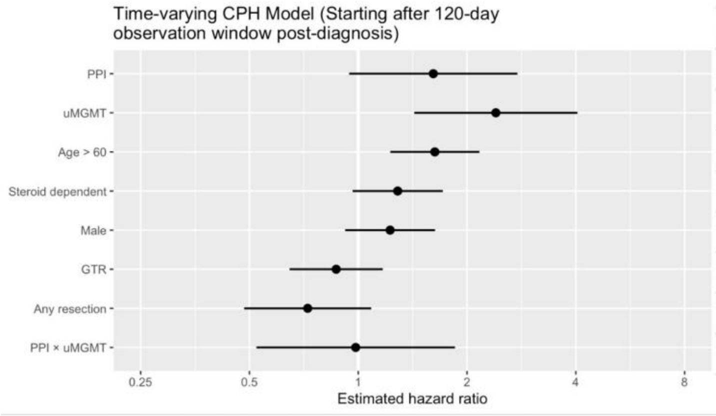


| HR | 95% confidence interval | Pr(>\|z\|) |
|---|---|---|
| 1.61 | (0.9439 to 2.758) | 0.080321 |
| 2.40 | (1.4285 to 4.044) | **0.000955** *** |
| 1.63 | (1.2262 to 2.165) | **0.000762** *** |
| 1.29 | (0.9644 to 1.714) | 0.086765 |
| 1.23 | (0.9199 to 1.632) | 0.164791 |
| 0.87 | (0.6457 to 1.17) | 0.354492 |
| 0.72 | (0.4833 to 1.086) | 0.119104 |
| 0.98 | (0.5227 to 1.852) | 0.960047 |

**Figure S4: Landmark analysis starting 120 days from diagnosis demonstrates similar hazard from PPI use.**

Landmark analysis of the time-varying CPH model starting 120 days from diagnosis maintains hazard with PPI use. Although the effect of PPI on hazard ratio is slightly outside of statistical significance (p = 0.08), data in the 120 days from diagnoses was discarded, reducing the statistical power.

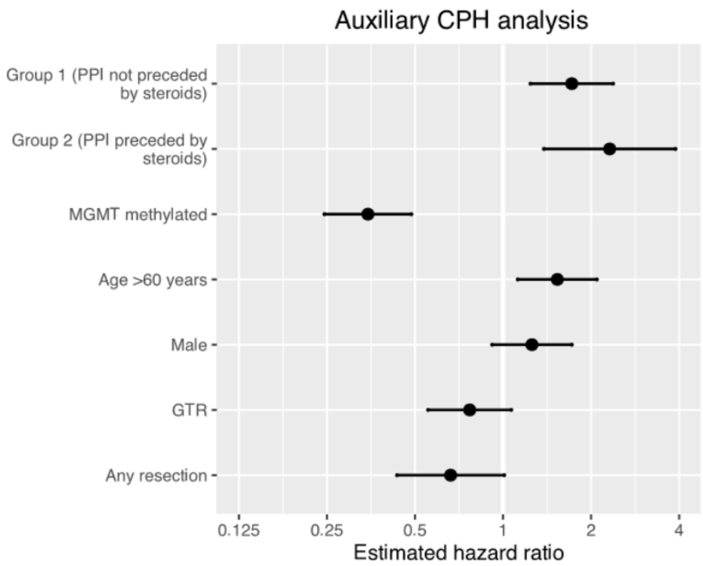


| HR | 95% confidence interval | Pr(>\|z\|) |
|---|---|---|
| 1.72 | (1.2405 to 2.3784) | **0.00112 **** |
| 2.31 | (1.3799 to 3.8823) | **0.00147 **** |
| 0.35 | (0.2453 to 0.4858) | **1.05e-09 ***** |
| 1.53 | (1.123 to 2.0917) | **0.00712 **** |
| 1.26 | (0.9179 to 1.7172) | 0.15446 |
| 0.77 | (0.5542 to 1.0668) | 0.11574 |
| 0.66 | (0.434 to 1.0089) | 0.05498 |

**Figure S5: Proton pump inhibitor use is hazardous to survival with and without proximal corticosteroid use**

Auxiliary time-varying Cox Proportional Hazard (CPH) model to consider impact of concurrent or temporally proximal use of corticosteroids and proton pump inhibitors (PPIs). '0' (baseline) if patients abstained from PPI use post-diagnosis, '1' if they initiated PPI use without concurrent corticosteroid use in the 15 days prior, and '2' if PPI initiation coincided with corticosteroid use within the preceding 15 days.
